# Improving the diagnosis of severe malaria in African children using platelet counts and plasma *Pf* HRP2 concentrations

**DOI:** 10.1101/2021.10.27.21265557

**Authors:** James A Watson, Sophie Uyoga, Perpetual Wanjiku, Johnstone Makale, Gideon M Nyutu, Neema Mturi, Elizabeth C George, Charles J Woodrow, Nicholas PJ Day, Philip Bejon, Robert O. Opoka, Arjen M Dondorp, Chandy C John, Kathryn Maitland, Thomas N Williams, Nicholas J White

**Affiliations:** Mahidol Oxford Tropical Medicine Research Unit, Faculty of Tropical Medicine, Mahidol University, Bangkok, Thailand; Centre for Tropical Medicine and Global Health, Nuffield Department of Medicine, University of Oxford, United Kingdom; KEMRI-Wellcome Trust Research Programme, Centre for Geographic Medicine Research-Coast, Kilifi 80108, Kenya; Medical Research Council Clinical Trials Unit, University College London, United Kingdom; Makerere University, Department of Paediatrics and Child Health, Kampala, Uganda; Indiana University, Department of Pediatrics, Indianapolis, Indiana, United States of America; Institute of Global Health Innovation, Department of Surgery and Cancer, Imperial College, London, United Kingdom

## Abstract

**Background:** Severe falciparum malaria is difficult to diagnose accurately in children in high transmission settings. Platelet counts and plasma concentrations of *P. falciparum* histidine-rich protein-2 (*Pf* HRP2) are potential biomarkers to increase diagnostic accuracy.

**Methods:** We fitted Bayesian latent class models to platelet counts and *Pf* HRP2 concentrations in 2,649 patients enrolled in four studies of severe illness in three countries (Bangladesh, Kenya, and Uganda). We estimated receiver operating characteristic curves and compared parasite densities, haematocrits, total white blood cell counts, blood culture positivity rates, and haemoglobin S genotypes (HbAS and HbSS) across the subgroups defined by the probabilistic models.

**Findings:** The platelet count and the plasma *Pf* HRP2 concentration have substantial diagnostic value in severe malaria. In severely ill patients with clinical features consistent with severe malaria, a combined platelet count ≤ 150,000 per *µ*L and a plasma *Pf* HRP2 concentration ≥ 1,000 ng/mL had an estimated sensitivity of 74% and specificity of 93% in identifying ‘true’ severe falciparum malaria. We estimate one third of African children enrolled in the two clinical studies of severe malaria had another cause of severe illness. Under the model, patients with severe malaria had higher parasite densities, lower haematocrits, lower rates of invasive bacterial disease, and a lower prevalence of both HbAS and HbSS than children misdiagnosed. Mortality in ‘true’ severe malaria was consistent across the African sites at ∼ 10%.

**Interpretation:** Studies of severe falciparum malaria in African children would be improved by including only patients with platelet counts ≤ 150,000 per *µ*L and plasma *Pf* HRP2 concentrations ≥ 1,000 ng/mL.

**Funding:** Wellcome

## Introduction

Severe falciparum malaria is defined clinically as vital organ dysfunction in presence of circulating *Plasmodium falciparum* parasites [1]. The primary goal of this definition is to identify severely ill patients rapidly and provide life-saving clinical management (notably parenteral artesunate treatment). This prioritises diagnostic sensitivity over specificity. In areas of moderate and high *P. falciparum* transmission, many apparently healthy children have malaria parasitaemia if their blood is examined either by light microscopy or a rapid diagnostic test. It follows that many hospitalised children will also be parasitaemic. As the major clinical features of severe malaria are not specific, it is difficult to differentiate clinically between severe falciparum malaria caused by extensive sequestration of malaria parasites in the microvasculature (i.e. ‘true’ severe malaria) and other causes of severe febrile illness accompanied by either coincidental asymptomatic parasitaemia or uncomplicated malaria [2, 3]. We have shown previously that complete blood counts (platelet counts and total white blood cell counts) provide substantial discriminating value in distinguishing true severe malaria from other causes of severe illness [4]. Approximately one third of a cohort of 2,220 Kenyan children diagnosed with severe malaria in a moderate transmission area were estimated to have had another cause of their severe illness [4]. The discriminative value of complete blood counts was validated using genetic polymorphism data in two different ways. First, the distribution of sickle trait (HbAS) was correlated very strongly with the estimated probability of severe malaria. The prevalence of HbAS was around five times higher in the subgroup of patients likely to have been mis-diagnosed compared to the subgroup with a likely correct diagnosis [4]. Second, we showed that genome-wide false discovery rates could be reduced substantially in a case-control whole-genome association study in which the output probability weights were used in a ‘data-tilting’ framework to adjust for patient mis-classification [4].

The diagnostic value of complete blood counts has great operational utility as blood counts are widely measured in routine practice at low cost. Moderate thrombocytopenia is a consistent feature of all human malaria infections [5, 6, 7], although the diagnostic utility of the platelet count has been debated [8, 9]. Platelets are activated in malaria infections and have increased turnover. In severe falciparum malaria there is endothelial activation with release of platelet aggregating activated high-multimeric von Willebrand Factor from specialized secretory vesicles in endothelial cells (the Weibel-Palade bodies). Platelets may also contribute to, and be consumed by, parasitised erythrocyte cytoadherence and autoagglutination [10, 11]. Severe thrombocytopenia is associated with increased mortality in severe malaria [9, 12, 13]. The total white blood cell count is also informative (although much less than thrombocytopenia), with a greater prognostic than diagnostic value (very high or very low white counts in severe disease are associated with high case fatality ratios) [7, 4]. In addition, total white counts vary according to age and ethnicity [14], confounding cross-study assessments.

The main pathophysiological process in severe falciparum malaria is the extensive sequestration of parasitised erythrocytes in the vascular beds of vital organs [15, 1]. These parasites, which cause potentially lethal pathology, have stopped circulating and are therefore not represented in the peripheral blood smear. In African children the peripheral parasite density is a poor indicator of disease severity and a poor diagnostic biomarker [16]. The parasite protein *Plasmodium falciparum* histidine-rich protein-2 (*Pf* HRP2), the basis for most rapid diagnostic tests, is liberated mainly at schizont rupture - and so the amount released is proportional to the extent of recent schizogony. Plasma *Pf* HRP2 is a much better discriminant of severe falciparum malaria than the peripheral blood parasite count [17, 18]. An example of how *Pf* HRP2 can help interpret clinical trial data comes from the large multi-centre AQUAMAT trial which was a randomised comparison of parenteral artesunate versus parenteral quinine in African children clinically diagnosed as having severe falciparum malaria. Overall, artesunate reduced the mortality by approximately 22% [19]. However, there was evidence for treatment effect heterogeneity across *Pf* HRP2 strata. In the subgroup of children in the highest tertile of *Pf* HRP2 artesunate reduced the mortality by one third (a similar proportion to that observed earlier in the SEAQUAMAT randomised comparison conducted in Southeast Asia [20]), whereas there was no substantial difference in mortality in the subgroup of children in the lowest tertile, i.e. the group with likely other causes of severe illness [18].

Defining optimal cutoff values for diagnostic biomarkers in severe malaria is difficult because in high-transmission areas there is no ‘gold-standard’ for diagnosis against which to calibrate thresholds. In the absence of a gold-standard, latent class models can be used to assess the sensitivity and specificity of threshold values for diagnostic indices [21]. Latent class models typically rely on having multiple biomarkers measured in the same individuals across different populations with varying disease prevalences, thus allowing for triangulation [22, 21]. We applied Bayesian parametric latent class models to admission platelet counts and plasma *Pf* HRP2 concentrations in combination, using data from four prospective studies of severe malaria or severe illness from Uganda, Kenya and Bangladesh, reflecting a range of *P. falciparum* transmission intensities from high to low, and thus a range of false diagnosis rates. The results suggest that the high rates of misdiagnosis could be reduced substantially by incorporating measurement of platelet counts and plasma *Pf* HRP2 concentrations in the diagnostic criteria.

## Results

### Platelet counts and *Pf* HRP2 concentrations in severe febrile illness

We pooled individual patient data from 2,649 severely ill African children and Asian adults, for whom platelet counts and measured plasma *Pf* HRP2 concentrations were available. The patients were from four separate studies in 3 countries:

- An observational study of severe falciparum malaria in Bangladesh (*n* = 172; all patients were clinically diagnosed with severe falciparum malaria).
- The Ugandan sites of the FEAST trial (*n* = 567, a randomised controlled trial of fluid resuscitation approaches in severe childhood illness not specific to severe malaria) [23].
- An observational study of cerebral malaria and severe malarial anaemia in Kampala, Uganda (*n* = 492) [24].
- A large cohort of children diagnosed with severe malaria in Kilifi, Kenya (*n* = 1, 418) [25].

Malaria transmission intensity is generally low in Bangladesh, moderate in Kilifi and Kampala, and high around the other Ugandan sites.

In total 27 patients (1%: 1 in Bangladesh, 8 in the FEAST sites in Uganda, and 18 from Kenya) had no detectable *Pf* HRP2 in the ELISA assay but had a parasite density >1,000 per *µ*L by microscopy. These could have either been assay errors or parasites with *Pf* HRP2/3 gene deletions.They were removed from the analysis, leaving a total of 2,622 samples. A summary of the patient characteristics in the analysed data set is shown in Table 1.

**Table 1:** Patient characteristics across the four studies. For age, parasite densities, platelet counts, total white blood cell counts and *Pf* HRP2 concentrations we show the median value and interquartile range. No haemoglobin S (HbS) genotyping was done for the patients from Bangladesh as HbS is absent from that population. *\**For FEAST, this excludes patients who had a negative RDT. IQR: inter-quartile range. HbAS: sickle trait; HbSS: homozygous sickle cell anaemia.

|  | Kilifi (Kenya) | Kampala (Uganda) | FEAST (Uganda) | Bangladesh |
| --- | --- | --- | --- | --- |
| *n* | 1,400 | 492 | 559 | 171 |
| Age (years, IQR) | 2.4 (1.4-3.7) | 3.3 (2.2-4.6) | 2.0 (1.2-3.3) | 30 (23-45) |
| Proportion parasite positive (%) | 100 | 100 | 59.4 | 100 |
| Parasite density* (per $\mu\text{L}$, IQR) | 69,824 (6,099-316,350) | 42,530 (10,635-198,540) | 37,600 (3,640-153,680) | 148,874 (23,550-348,540) |
| Platelet count ($10^3/\mu\text{L}$, IQR) | 111 (64-215) | 96 (49-170) | 165 (75-326) | 50 (27-139) |
| *Pf*HRP2 (ng/mL, IQR) | 2,207 (419-5,072) | 1,838 (588-4,097) | 175 (0-1,953) | 2,667 (1,083-6,128) |
| White blood cell count ($10^3/\mu\text{L}$, IQR) | 12.6 (8.9-19) | 10.4 (7.5-15.3) | 12.0 (8.4-18.7) | 9.0 (6.9-11.0) |
| Mortality (%) | 11.1 | 6.7 | 11.4 | 26.9 |
| HbAS (*n*, %) | 41 (2.9) | 4 (0.8) | 46 (8.2) | - |
| HbSS (*n*, %) | 7 (0.5) | 23 (4.7) | 21 (3.8) | - |

Log_10_-transformed platelet counts and log_10_-transformed *Pf* HRP2 concentrations were strongly inversely correlated in both the studies of severely ill African children (*ρ* =-0.54, *p* = 10^−186^), and in the Bangladeshi adults with severe malaria (*ρ* = -0.36, *p* = 10^−5^). Patients with platelet counts in the normal range (>150,000 per *µ*L) generally had low to non-measurable plasma *Pf* HRP2 concentrations (median concentration was 269 ng/mL, IQR: 24 to 1,043), while patients with thromobocytopenia (≤ 150,000 per *µ*L) had a median *Pf* HRP2 concentration of 3,031 ng/mL (IQR: 1,261 to 6,035).

### Discriminative value of platelet counts and plasma *Pf* HRP2 in patients diagnosed with severe malaria

We first assessed the value of platelet counts and plasma *Pf* HRP2 concentrations in patients who were diagnosed with severe malaria (a total of *n*=2,063; this excluded patients from the FEAST study which explicitly included non malarial causes of severe febrile illness). We fitted a two component parametric Bayesian latent class model (with the two latent classes representing ‘severe malaria’ and ‘not severe malaria’) using the log_10_ transformed biomarkers (see Methods for description of the informative priors used and the key assumptions). Figure 1 shows the model estimated sensitivities and specificities for all cutoff values of the platelet count and the *Pf* HRP2 concentration. As expected, the platelet count and the *Pf* HRP2 concentration both had high discriminative value (illustrated by the receiver operating characteristic (ROC) curves). Overall, for thresholds giving the same sensitivity, the plasma *Pf* HRP2 concentration had a higher specificity. For example, in these populations, a lower limit of 1000 ng/mL for the *Pf* HRP2 concentration had an estimated sensitivity of 87% and a specificity of 83%. In comparison, an upper limit for the platelet count of 150,000 per *µ*L had an estimated sensitivity of 83% but a specificity of 71%.

**Figure 1:**
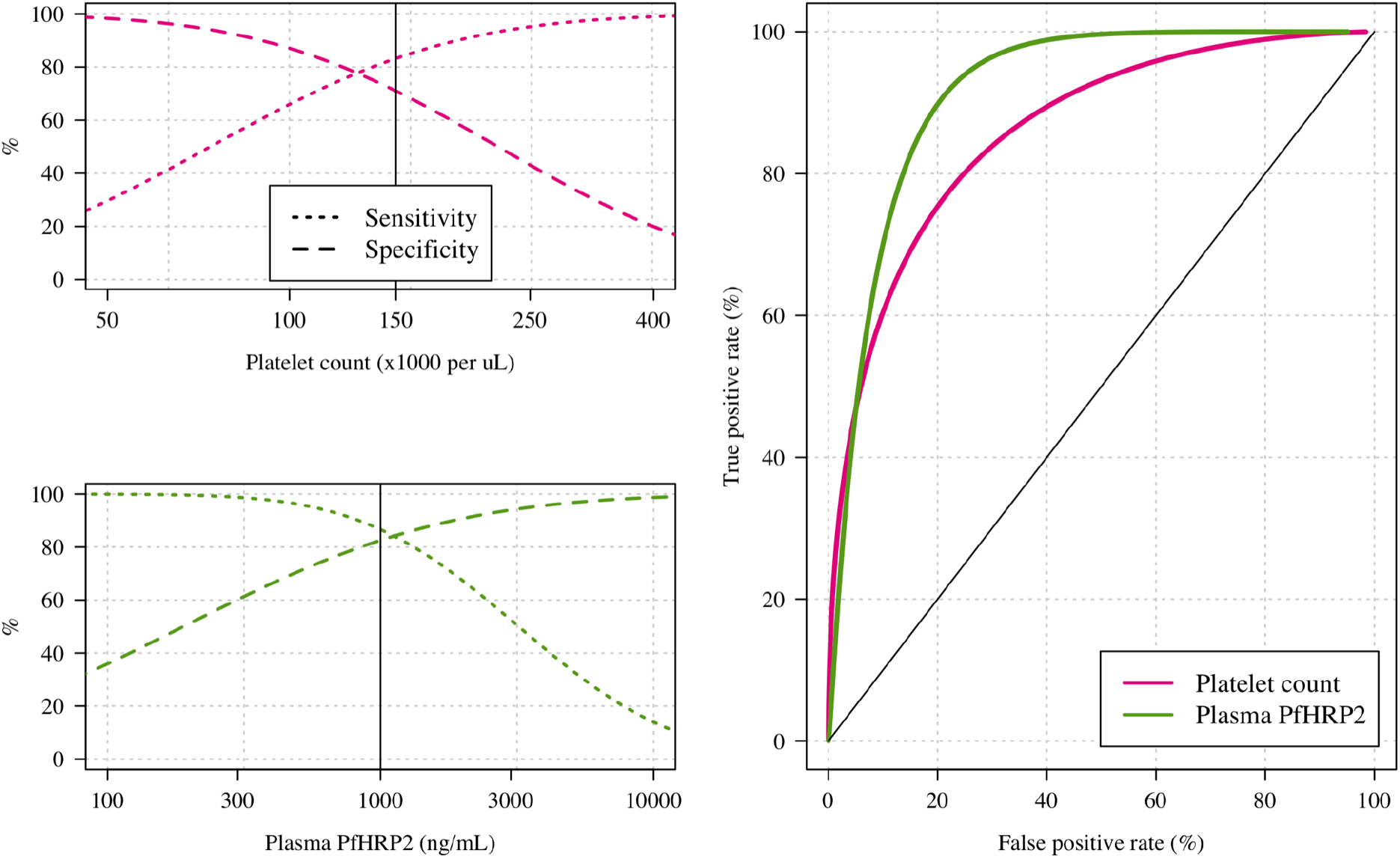
The diagnostic value of platelet counts (pink) and plasma *Pf* HRP2 concentrations (green) estimated using data from 2,063 patients diagnosed with severe falciparum malaria in three studies. Sensitivities (dotted lines) and specificities (dashed lines) are estimated under a Bayesian parametric two-component latent class model which assumes the same sensitivity and specificity for each biomarker across the three studies (African children: Kilifi, Kenya and Kamapala, Uganda; Asian adults: Bangladesh). For the platelet count, thresholds correspond to upper limits, whereas for the *Pf* HRP2 concentration the thresholds correspond to lower limits.

A series of sensitivity analyses (using non-informative priors, different choices for the parameteric model, or using only data from the two studies of African children with severe malaria) showed near identical results (Figure S1).

### Joint diagnostic thresholds for platelet counts and *Pf* HRP2

For clinical and epidemiological studies, and in contrast to clinical practice, specificity is usually more important that sensitivity. We used both biomarkers in combination to improve the precision of the definition of severe malaria, in order to achieve low false positive rates. Figure 2) shows the false positive and true positive rates when using both the platelet count and the plasma *Pf* HRP2. Multiple combinations of the platelet count and *Pf* HRP2 concentration have approximately the same operating characteristics in diagnosis, so the optimal choice is subjective and can be made based on operational simplicity. In severe illness with clinical features consistent with severe malaria, platelet counts ≤ 150,000 per *µ*L and plasma *Pf* HRP2 concentrations ≥ 1000 ng/mL in combination have an estimated diagnostic specificity of 93% and a sensitivity of 74%. If this definition was applied to a hospital cohort with a prevalence of severe malaria of 60% (approximately the prevalence estimated for the Kenyan cohort of children with clinically diagnosed severe malaria), we would expect over 94% of the resulting population identified by these two biomarkers to have ‘true’ severe malaria (positive predictive value).

**Figure 2:**
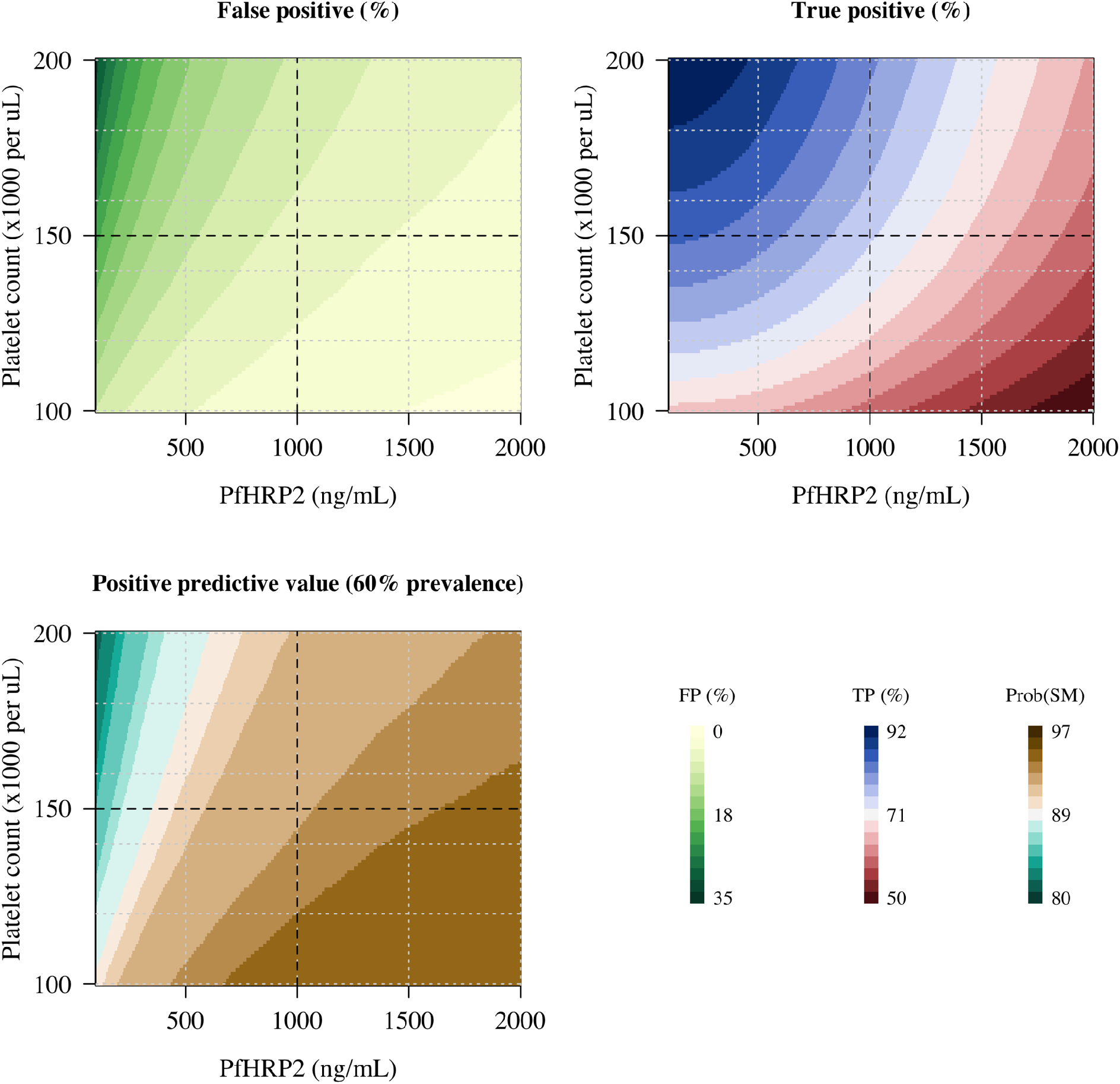
The estimated operating characteristics for the platelet count and the plasma *Pf* HRP2 concentration when used in combination to diagnose severe falciparum malaria. The top two panels show the false positive and true positive rates as a function of the joint platelet count and *Pf* HRP2 thresholds. The bottom left panel shows the corresponding positive predictive value assuming a 60% prevalence of severe malaria in target population (hospitalised children with clinical features consistent with severe malaria). TP: true positive rate; FP: false positive rate. Prob(SM): posterior probability of severe malaria under the model.

### Estimating the probability of severe malaria using platelet counts and *Pf* HRP2

We fitted a three component parametric Bayesian latent class model to all available platelet count and *Pf* HRP2 concentration data from patients from the four studies of severe febrile illness (a total of *n*=2,622) in order to estimate the individual patient probabilities that severe malaria was the true cause of their severe illness, and to calculate the prevalences of ‘true’ severe malaria among those diagnosed. Under this model, we estimated that the prevalence of true severe malaria was 96.1% (95% CI: 91.0-98.8) in Bangladeshi adults; 36.6% (95% CI: 31.4-41.9) in the Ugandan children enrolled in the FEAST trial (FEAST intentionally enrolled severely ill children with and without malaria, although 66% of all patients had a clinical diagnosis of severe malaria [23]); 73.5% (95% CI: 66.6-79.4) in the children enrolled in Kampala, Uganda [24]; and 65.9% (95% CI: 61.0-70.2) in the children diagnosed with severe malaria in Kilifi, Kenya.

Figure 3 shows scatter plots of the platelet counts versus the plasma *Pf* HRP2 concentrations, coloured by the probability that the patient had severe malaria and grouped by study (dark blue: high probability of severe malaria; dark red: low probability of severe malaria). The model estimated a geometric mean platelet count in severe malaria across the four studies of 74,000 per *µ*L (95% of patients are predicted to have platelet counts between 17,000 and 312,000) and a geometric mean *Pf* HRP2 of 3,135 ng/mL (95% prediction interval: 402 to 24,452). The model-based probabilities of severe malaria were highly concordant with our previously published model which used platelet counts and total white blood cell counts (*ρ* =0.63, Figure S2) [4].

**Figure 3:**
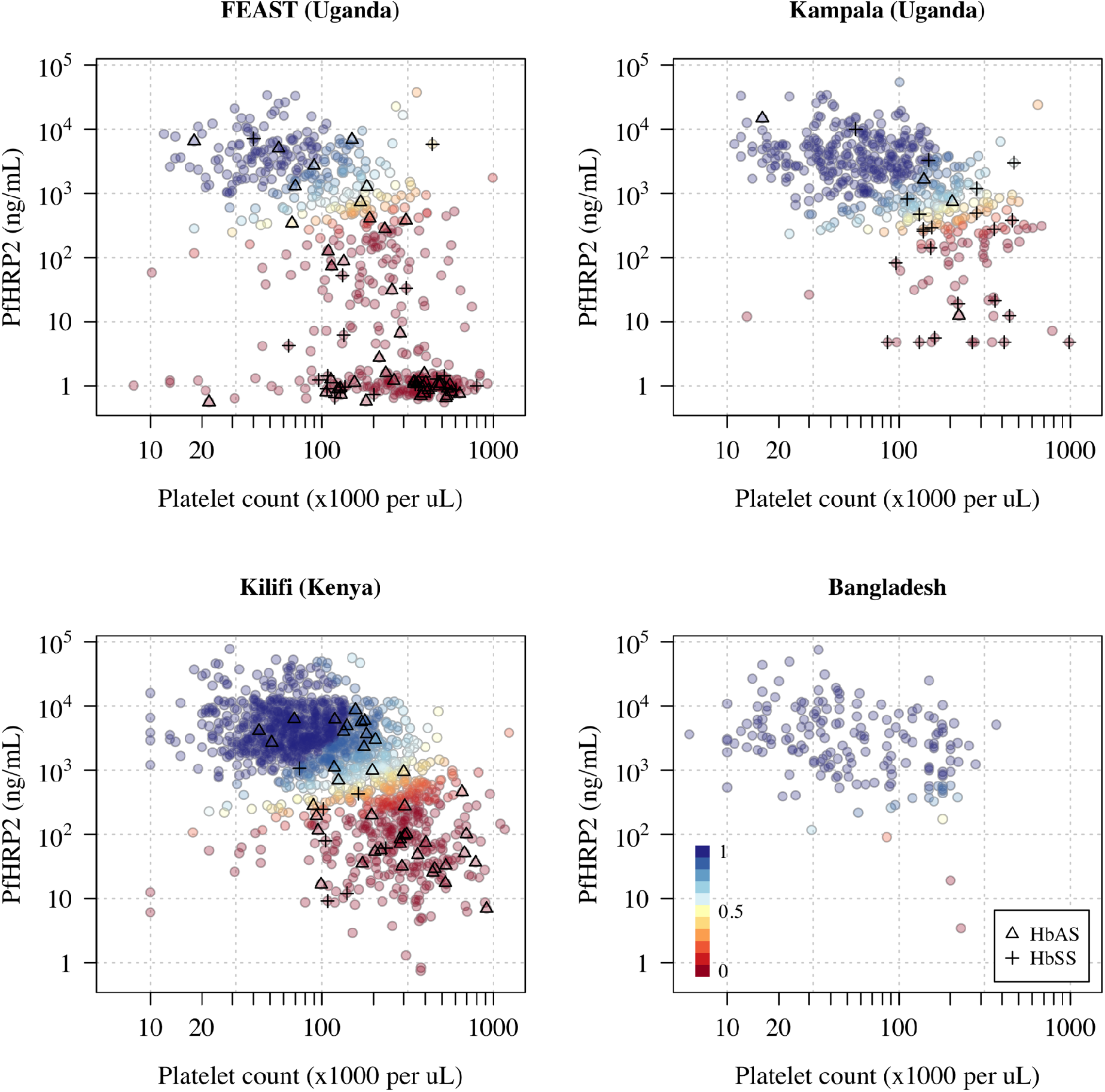
Probabilistic model of severe falciparum malaria using platelet counts and plasma *Pf* HRP2 concentrations in 2,622 severely ill patients based on a Bayesian parametric latent class model with three latent classes (a severe malaria class and two ‘not severe malaria’ classes). The colours correspond to the probability of severe malaria under the model (dark blue: high probability; dark red: low probability). Triangles show the HbAS individuals; crosses show the HbSS individuals. In order to show data points with non-measurable plasma *Pf* HRP2, non-measurable concentrations were set to 1 ng/mL ± random jitter on the log_10_ scale (approximately half the lower limit of quantification of the assay).

We compared the estimated false diagnosis rates for cerebral malaria versus severe malaria without coma, and for severe malarial anaemia versus severe malaria without severe anaemia (where severe anaemia is defined as a haematocrit ≤ 15%) in the two cohorts of children clinically diagnosed with severe malaria. In the Kenyan cohort, the estimated false diagnosis rate was slightly higher in the cerebral malaria group relative to the non-cerebral malaria groups: 36% of patients with coma, and 26% of patients without coma were classified as “falsely diagnosed” with severe malaria (*p* =0.001). For the subgroup with severe anaemia, the false diagnosis rate was 15%, compared to 36% in the subgroup without severe anaemia. In the Ugandan cohort, both of these trends were reversed. A false diagnosis of severe malaria was estimated for only 12% of patients with coma, but for 40% of patients without coma. For severe anaemia, these proportions were 36% and 15%, respectively (the Ugandan cohort only recruited patients with either severe malarial anaemia or patients with cerebral malaria).

Mortality rates varied substantially as a function of the estimated probability of severe malaria and across the studies (Figure 4). In African children with a high probability of having severe malaria, the mean mortality was consistently around 10% for the three studies included (Figure 4). Apart from the Bangladeshi adults, all patients were treated with initial intravenous quinine. Mortality in severe malaria in adults from Bangladesh was substantially higher (∼30%). In Kenyan children, the mortality in the mis-classified group of patients was higher than in the correctly classified patients (14% versus 10%), whereas in the Ugandan study the trend was reversed (1% versus 10%). This is largely explained by the study populations: the group with a low probability of having severe malaria in the Ugandan study was predominantly composed of patients with severe anaemia without other features of severity.

**Figure 4:**
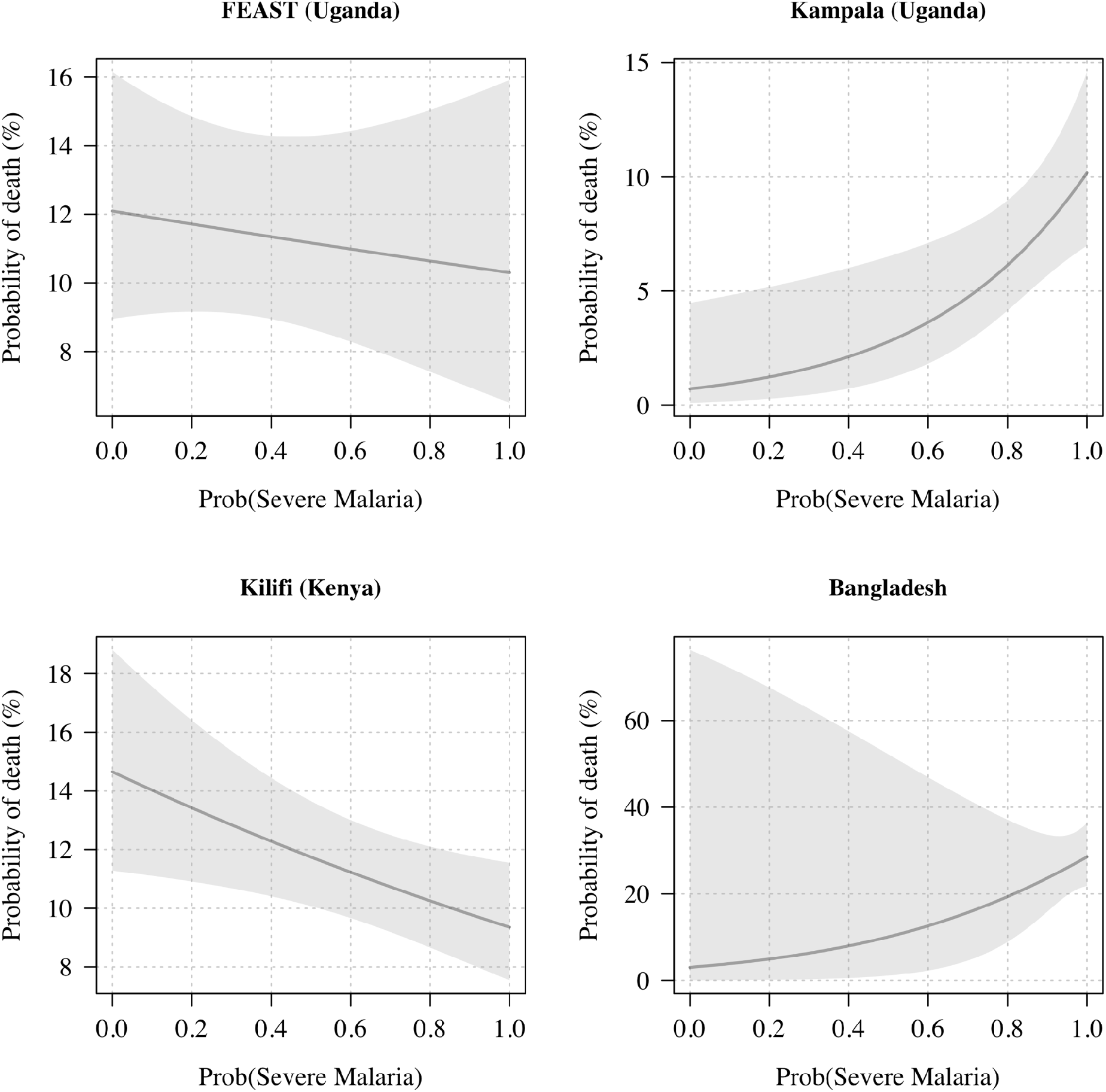
Mortality as a function of the probability of having severe malaria under the Bayesian latent class model (based on platelet counts and *Pf* HRP2 concentrations). The lines (shaded areas) show mean (95% confidence intervals) mortality estimates from logistic regression fits.

### Relationship with other admission variables

We explored the relationship between the model estimated probability of severe malaria and the admission parasite densities, admission haematocrits, the total white blood cell counts, and the blood culture positivity rates (i.e. cultures growing a likely pathogen). In the three African studies, parasite densities were between 12 and 16 times higher in patients with a high probability of severe malaria versus those with a low probability of severe malaria (Figure 5). After adjusting for study differences, the parasite densities were estimated to be 12.8-fold higher (95% C.I 9.9-16.5, p=10^−79^) in severe malaria versus not severe malaria.

**Figure 5:**
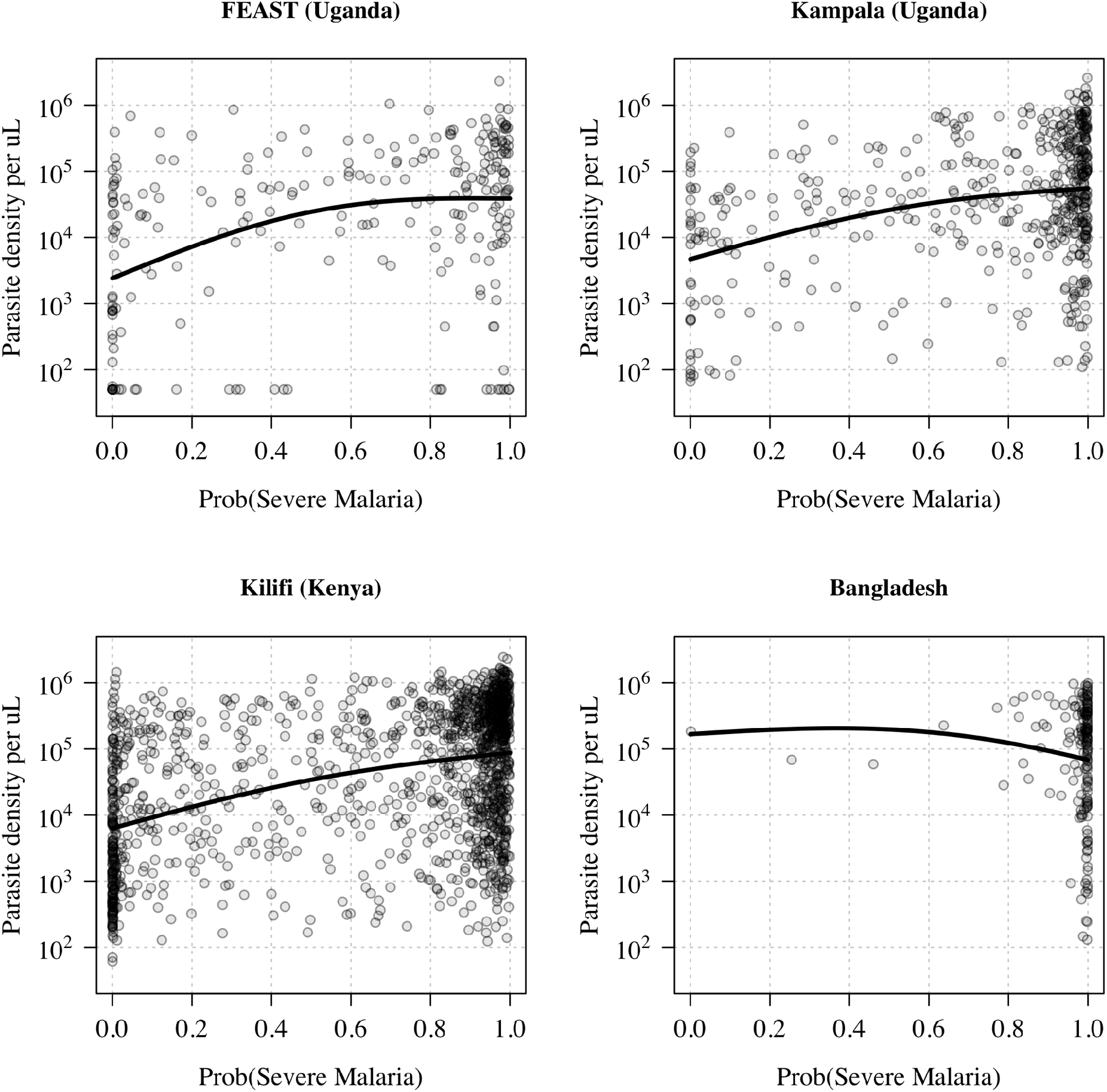
Admission parasite densities as a function of the probability of severe malaria under the Bayesian latent class model. Data from the FEAST trial includes only the malaria RDT positive patients. The thick lines show the additive linear model fit (spline based).

The admission haematocrits were also highly correlated with the model estimated probability of severe malaria (Figure 6). In the three African studies, children with a high probability of having severe malaria had median admission haematocrits between 16 and 20% (FEAST: 19%; Kampala: 16%; Kilifi: 20%). The haematocrit distributions in this group were unimodal (Figure S4). In contrast, the haematocrit distributions in patients with low probabilities of having severe malaria (<0.2) were strongly bi-modal, with the majority of patients having higher haematocrits (the median haematocrits in this group were: FEAST: 30%; Kampala: 12%; Kilifi: 28%), but a substantial minority had low haematocrits of around 10%. The few Bangladeshi adults with a low probability of severe malaria also had higher haematocrits.

**Figure 6:**
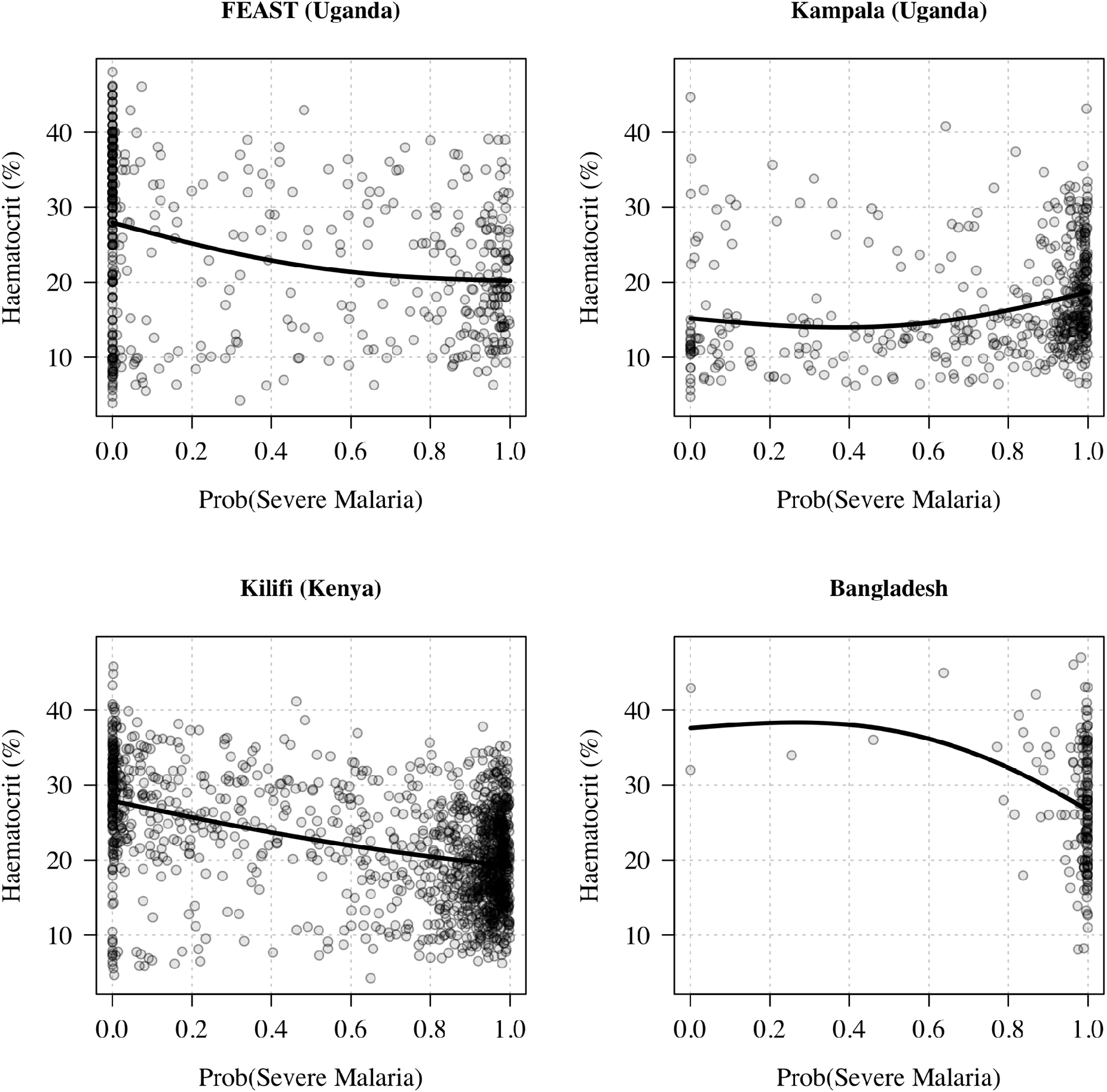
Admission haematocrits as a function of the probability of severe malaria under the Bayesian latent class model (model probabilities based on the platelet counts and the plasma *Pf* HRP2 concentrations).

Blood cultures were done for all 1400 Kenyan children and for 298 out of 332 (90%) Ugandan children in the FEAST study who had malaria parasitaemia on admission. Overall 51 and 35 patients respectively had positive blood cultures. The probability of having severe malaria was highly predictive of the blood culture result, with an adjusted odds-ratio of 0.43 (95% CI: 0.27-0.66; p=0.0002) for a positive culture in patients likely to have severe malaria versus those unlikely to have severe malaria (after adjustment for study). This difference in blood culture positivity rates was also reflected in the total white blood cell counts (Figure S3). Across the four studies, after adjustment for age, in patients likely to have severe malaria (probability > 0.5) compared to those unlikely to have severe malaria (probability < 0.5) the odds-ratio for having a total white count > 15,000 per *µ*L was 0.50 (95% CI: 0.46-0.56; p=10^−12^).

### Gene-dose relationship for haemoglobin S and severe malaria

In the three studies in African children, the prevalence of both HbAS and HbSS were strongly inversely correlated with the model estimated probability of severe malaria (Figure 3). Pooling the three African studies and adjusting for differences in the estimated prevalence of severe malaria, the odds-ratio for being classified as severe malaria (probability > 0.5) for HbAS patients relative to HbAA patients was 0.24 (95% CI: 0.15 to 0.40, *p* = 10^−8^), and for HbSS relative to HbAA the odds-ratio was 0.08 (95% CI: 0.04 to 0.18, *p* = 10^−10^). Under an additive model of association, each additional haemoglobin S (HbS) allele was associated with an odds-ratio for severe malaria of 0.27 (95% CI: 0.19 to 0.37, *p* = 10^−16^). This association between HbS genotypes and the biomarker-based severe malaria classification was highly concordant across the three studies.

## Discussion

The diagnosis of severe malaria in African children is imprecise [3, 26, 2]. This is because it is difficult to distinguish clinically between severe illness caused by malaria from severe illness with incidental asymptomatic or uncomplicated malaria since they share many clinical characteristics. This is a substantial problem for studies of one of the most important life threatening infections in childhood. It dilutes and distorts the results of epidemiology [27], pathophysiology [2], genetic association [4] and therapeutic investigations [18]. In areas of moderate and high levels of malaria transmission asymptomatic parasitaemia is very common. At any time a high proportion of children have detectable malaria parasitaemia. Malaria parasitaemia is therefore a sensitive but not specific indicator that malaria is the cause of illness [26]. Other simple laboratory tests provide valuable diagnostic information. Thrombocytopenia is a common feature of all symptomatic malarias [5, 6, 7]. A low platelet count therefore supports (but does not prove) attributing malaria as the cause of illness. We estimate that less than one in five patients with severe malaria will have platelet counts in the normal range (>150,000 per *µ*L). This analysis of data from large prospective studies of severe illness in African children shows that measurement of platelet counts and the plasma concentration of the parasite protein *Pf* HRP2 can together substantially improve the specificity of a diagnosis of severe falciparum malaria.

A key strength of this work is that we can validate the discriminant power of the platelet count and the plasma *Pf* HRP2 concentration by comparing the prevalence of HbS genotypes (HbAS and HbSS) and the blood culture positivity rates across in the inferred subgroups. HbAS is the genotype that provides the strongest known protection against severe falciparum malaria [28, 29]. The prevalence of HbAS was four times lower in children considered likely to have severe malaria compared with those considered less likely to have severe malaria. It is important to note that HbAS also protects against complications of malaria, such as an increased risk of bacterial infections, so the HbAS prevalence in children with presumed bacterial infections is still expected to be lower than that in the healthy population [30]. Recent work suggests that the protective effect of HbAS may vary according to parasite genotype [31]. It may be that the few HbAS individuals who have “true” severe malaria have parasite genotypes that can evade the HbAS defence mechanisms. Future work will assess the relationship between these probabilistic classifications and the parasite genotypes. Whereas people with the AS genotype are essentially haematologicaly normal, those with homozygous SS suffer from sickle cell disease. This causes anaemia, and may also cause thrombocytopenia and leukocytosis. This could confound the use of full blood count data in the probabilistic assessment of severe malaria. However plasma *Pf* HRP2 (or other parasite biomass indicators) should not be affected by sickle cell disease. In this study there was strong evidence for an additive effect between the number of HbS alleles and a decreased probability of severe malaria under the platelet/HRP2 model. This suggests that HbSS is strongly protective against a high parasite biomass [32]. However as sickle crises may be severe, and they are often triggered by infections, it is possible a low parasite biomass could trigger severe events. Whether or not this should be described as ‘severe malaria’ is a semantic question [33]. A recent in-depth analysis of the Ugandan cohort from Kampala showed that the HbSS children considered to have severe malaria had a lower parasite biomass but higher levels of endothelial dysregulation [32]. The main differential diagnosis in suspected severe malaria is bacterial sepsis. The relationship is complicated as severe malaria does predispose to bacterial sepsis [34]. Blood culture has low sensitivity, however we observed a three fold increase in positivity rates in the patients likely have been mis-diagnosed compared to those with a likely correct diagnosis.

The main limitation of this study is the absence of a gold-standard diagnosis, and thus the reliance on parametric latent class models. The application of latent class models to multiple populations with varying disease prevalence provides a powerful framework for estimating the diagnostic accuracy of imperfect tests, but the model outputs rely on the validity of key assumptions, notably the underlying parametric models used for the biomarker distributions. We used log-normal distributions for the platelet count and plasma *Pf* HRP2 concentration in both ‘true’ severe malaria and ‘not severe malaria’. Visual data plots suggest that this is a good approximation of the true underlying distributions. Each individual biomarker has its own limitations. Thrombocytopenia can be caused by other infections (e.g. severe arbovirus infection) and may occur in sepsis, although it is much less prevalent in sepsis than malaria [7]. The main limitation of plasma *Pf* HRP2 as a marker of total parasite biomass is that its plasma clearance is slow relative to the parasite life-cycle (3-4 days) [17] and it requires specialist measurement (although rapid tests can be applied to plasma [35] and modifications are under development). As plasma *Pf* HRP2 accumulates each asexual parasite cycle, an individual with a sustained low parasite multiplication rate at high parasite densities can have the same *Pf* HRP2 concentration as an individual with a fulminant infection and a high sequestered biomass [17]. In addition mutations in the *Pf* HRP2 gene causing changes in antigenicity may affect immunoassays [36, 37].

In these very large prospective series of patients hospitalised with a diagnosis of severe malaria, combining platelet counts and plasma *Pf* HRP2 concentrations provided good discrimination between “true” severe falciparum malaria and other severe illnesses (likely to be bacteraemia in many of the cases) with concomitant incidental malaria. A major strength of this study is in combining two measures which are mechanistically distinct (platelet counts are measuring the host response to acute malaria illness; *Pf* HRP2 is measuring the parasite biomass). We show that in combination these biomarkers provide a high level of discrimination between patients who were likely to have had true severe malaria and those with a different illness aetiology. This proportion ranged from one third in the FEAST study which intentionally studied children with both severe malaria and other severe illnesses (likely mainly sepsis) requiring fluid resuscitation to >90% in Bangladesh -a low transmission area where the diagnosis of falciparum malaria as the cause of illness is highly specific. Overall it suggests that approximately one third of African children diagnosed with severe malaria have another cause of their severe illness [4]. Studies of severe falciparum malaria would be improved by including only children with platelet counts of ≤ 150,000 per *µ*L and plasma *Pf* HRP2 concentrations of ≥ 1,000 ng/mL.

## Methods

### Data

All the clinical studies were prospective studies of severe illness and had appropriate ethical approval. In all four studies plasma *Pf* HRP2 levels were quantitated using the same previously published methodology [18].

### Bangladesh

We included data from observational studies in severe falciparum malaria conducted by the Mahidol Oxford Tropical Medicine Research Unit in Bangladesh between 2003 and 2019. These pooled data have been described previously [38]. Platelet counts and *Pf* HRP2 concentrations were jointly measured in a total of 172 patients. The majority of these patients received intravenous artesunate.

### Kilifi (Kenya)

The Kenyan case-control cohort has been described in detail previously [25]. Severe malaria cases consisted of all children aged <14 years who were admitted with clinical features of severe falciparum malaria to the high dependency ward of Kilifi County Hospital between June 11th 1999 and June 12th 2008. Severe malaria was defined as a positive blood-film for *P. falciparum* along with: prostration (Blantyre Coma Score of 3 or 4); cerebral malaria (Blantyre Coma Score of <3); respiratory distress (abnormally deep breathing); severe anaemia (haemoglobin < 5 g/dL). Cases and controls were genotyped for the rs334 SNP (HbS) using DNA extracted from fresh or frozen samples of whole blood as described in detail previously [25, 39]. The standard of care antimalarial treatment during this period was intravenous quinine.

### FEAST (Uganda)

FEAST was a multicentre randomised controlled trial comparing fluid boluses for severely ill children with shock (*n* = 3, 161) that was not specific to severe malaria [23]. Platelet counts and plasma *Pf* HRP2 concentrations were measured in 502 children in the Ugandan sites (Mulago National Referral Hospital, Mbale and Soroti Regional Referral Hospitals and St Mary’s Hospital, Lacor). The standard of care antimalarial treatment during this period was intravenous quinine.

### Kampala (Uganda)

The trial by Brand *et al* was an observational study of cerebral malaria and severe malarial anaemia in Mulago Hospital, Kampala, Uganda [24]. Children were enrolled if they were between 18 months and 12 years of age. Cerebral malaria was defined as coma (Blantyre Coma Score of <3 or Glasgow Coma Score <8) in presence of parasites on a blood smear. Severe malarial anaemia was defined as presence of *Plasmodium falciparum* on blood smear in children with hemoglobin level ≤5 g/dL. The standard of care antimalarial treatment during this study was intravenous quinine.

### Statistical analysis

We fitted a series of Bayesian parametric latent class models to the available biomarker data [21]. The key assumptions of the main models are summarised as follows:

1. The marginal distribution of each biomarker in each latent class is log-normal.
2. The marginal distribution of each biomarker in each latent class is the same across the different studies and countries.
3. Informative Bayesian priors on all parameters.

Sensitivity analyses relaxed assumptions 1-3 (using bivariate t-distributions, only using data from African children, and using weakly informative priors). We compared models with and without correlation between the biomarkers within each latent class. The models with correlation performed better and were more conservative.

The first set of models used data from the three severe malaria studies only (Kenyan children from Kilifi, Ugandan children from Kampala, and Bangladeshi adults). For each biomarker, the marginal distribution in each latent class (severe malaria versus not severe malaria) was assumed to be log-normal with the following informative priors (means and standard deviations are given on the log_10_ scale):

- The mean platelet count: *N* (log_10_ 75, 0.1) for severe malaria and *N* (log_10_ 200, 0.1) for not severe malaria.
- The mean *Pf* HRP2: *N* (log_10_ 3000, 0.2) for severe malaria and *N* (log_10_ 250, 0.2) for not severe malaria.
- The standard deviation on the log_10_ scale for each log-normal distribution was given a exponential prior with rate parameter set to 2 (i.e. a mean standard deviation of 0.5 on the log_10_ scale)

We used informative beta priors for the prevalence of severe malaria in the three studies: Beta(19,1) for Bangladesh; Beta(14,6) for Kilifi; Beta(14,6) for Kampala.

To assess the robustness of the model outputs, we performed three sensitivity analyses: (i) changing the parametric form to a bivariate student t-distribution with 10 degrees of freedom (robustness to assumption 1); (ii) by fitting the model to data only from the studies in African children with severe malaria (Kampala and Kilifi; robustness to assumption 2); (iii) by using weakly informative priors (see code).

For the second set of models, we included data from the FEAST trial [23]. FEAST intentionally enrolled severely ill patients with and without malaria, thus there is a subset of patients who are clearly distinct from all patients in the studies of severe malaria (this subset is characterised on average by a negative malaria blood slide, no measurable plasma *Pf* HPR2 and a normal platelet count). The latent class model needed to include a third component in order to fit the data. Under this model, the biomarker distributions were also assumed to be log-normal in the severe malaria class, but were a mixture of two log-normal distributions for the ‘not severe malaria’ class.

We used the same priors for the second set of models with the addition of the following priors for the third additional component (which can be summarised as severe illness with no evidence of acute malarial infection):

- The mean platelet count: *N* (log_10_ 300, 0.1) for not severe malaria.
- The mean *Pf* HRP2: *N* (log_10_ 2, 0.2) for not severe malaria.

Dirichlet priors were used for the prevalence parameters for each latent class: Dirichlet(19,1,0.1) for the Bangladesh study (with hyperparameters corresponding to the severe malaria and the two not severe malaria classes, respectively); Dirichlet(3,3,3) for the FEAST trial; Dirichlet(14,7,1) for the Kampala and Kilifi studies.

In order for patients with non-measurable plasma *Pf* HRP2 concentrations to be included in this analysis, we set all non-measurable concentrations to 1 ng/mL (approximately half the lower limit of detection of the ELISA assay). Patients who had non-measurable plasma *Pf* HRP2 concentrations but parasite densities above 1,000 per *µ*L were excluded from the analysis.

All posterior distributions were estimated using Monte Carlo methods. All models were implemented in the *rstan* language [40]. For each model we ran 4 independent chains for 2000 iterations, discarding half for burn-in. Convergence was checked by assessing traceplots. Convergence problems due to ‘label switching’ were avoided by using parameter constraints: the mean value of each likelihood distribution was increasing (for the platelet count) or decreasing (for the *Pf* HRP2 and parasite density). This uses the *ordered* parameter class in *stan*. The *stan* language does not support discrete parameters, however posterior distributions of latent class models can be sampled by using the *logsumexp* trick, computing the marginal likelihood over all possible class combinations.

## Data Availability

A minimal clinical dataset is available online via the following github repository:
https://github.com/jwatowatson/SevereMalariaDiagnosis.

## Code and data availability

All code in the form of an RMarkdown document along with a minimal clinical dataset is available online via the following github repository: https://github.com/jwatowatson/SevereMalariaDiagnosis. A static version can be found at https://doi.org/10.5281/zenodo.5602924

## Acknowledgements

This research was funded by Wellcome. A CC BY or equivalent licence is applied to the author accepted manuscript arising from this submission, in accordance with the grant’s open access conditions.

This work was done as part of SMAART (Severe Malaria Africa – A consortium for Research and Trials) funded by a Wellcome Collaborative Award in Science grant (209265/Z/17/Z) held in part by KM, NPJD and AMD.

JAW is a Sir Henry Dale Fellow jointly funded by the Wellcome Trust and the Royal Society (223253/Z/21/Z). TNW and NJW are senior and principal research fellows respectively funded by the Wellcome Trust (202800/Z/16/Z and 093956/Z/10/C, respectively). ECG acknowledges funding from a core grant to the MRC CTU at UCL from the MRC (MC_UU_12023/26). CCJ and ROO acknowledge grant R01 NS055349 from the National Institutes for Neurologic Disorders and Stroke. The FEAST trial was supported by a grant (G0801439) from the Medical Research Council, United Kingdom provided through the (MRC) DFID concordat. KM and ECG were supported by this grant. This paper is published with permission from the Director of the Kenya Medical Research Institute (KEMRI).

## Author contributions

JAW conceived the study, designed the experiments, analysed the data and wrote the first draft of the paper. NJW, TNW, KM supervised the work. SU, PW, JM, GMN, NM, ECG, CW, NPJD, PB, ROO, AMD, CJ, KM, TNW, NJW contributed data from the clinical studies. All authors read and revised the paper.

---

**Figure S1:**
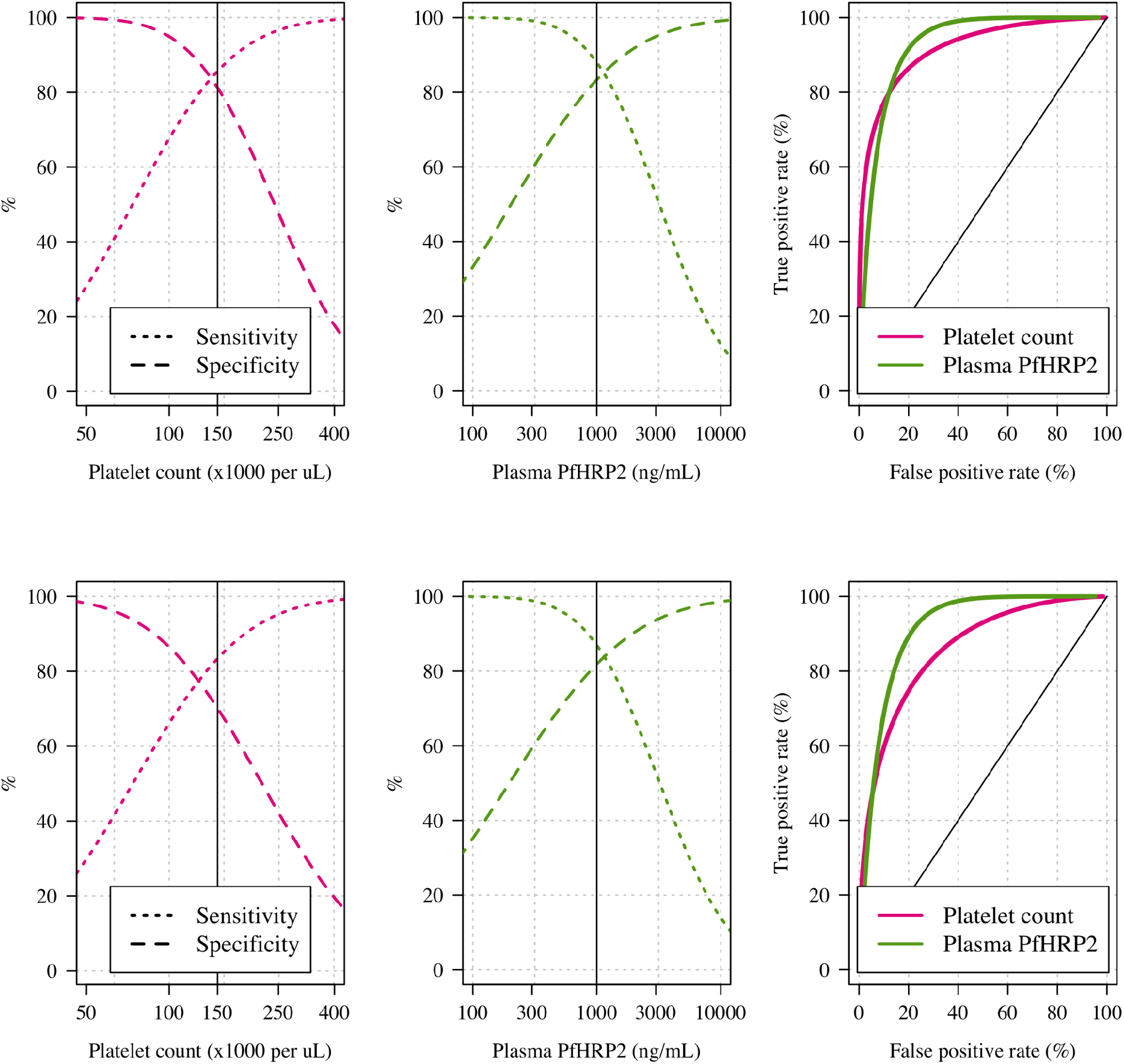
Receiver operating characteristics for the two main sensitivity analyses performed. The top row shows the same panels as in the main Figure 1 but with a model using bivariate t-distributions instead of bivariate normal distributions. The bottom row shows the same panels but with weakly informative priors (see code). When using a bivariate t-distribution, the model estimates a higher specificity for the platelet count but from visual inspection of the patients for whom the posterior probability changes considerably, we decided that this is most likely to be wrong, reflecting prior beliefs that the plasma HRP2 is more specific (thrombocytopenia can be caused by other diseases or conditions such as sickle cell anaemia).

**Figure S2:**
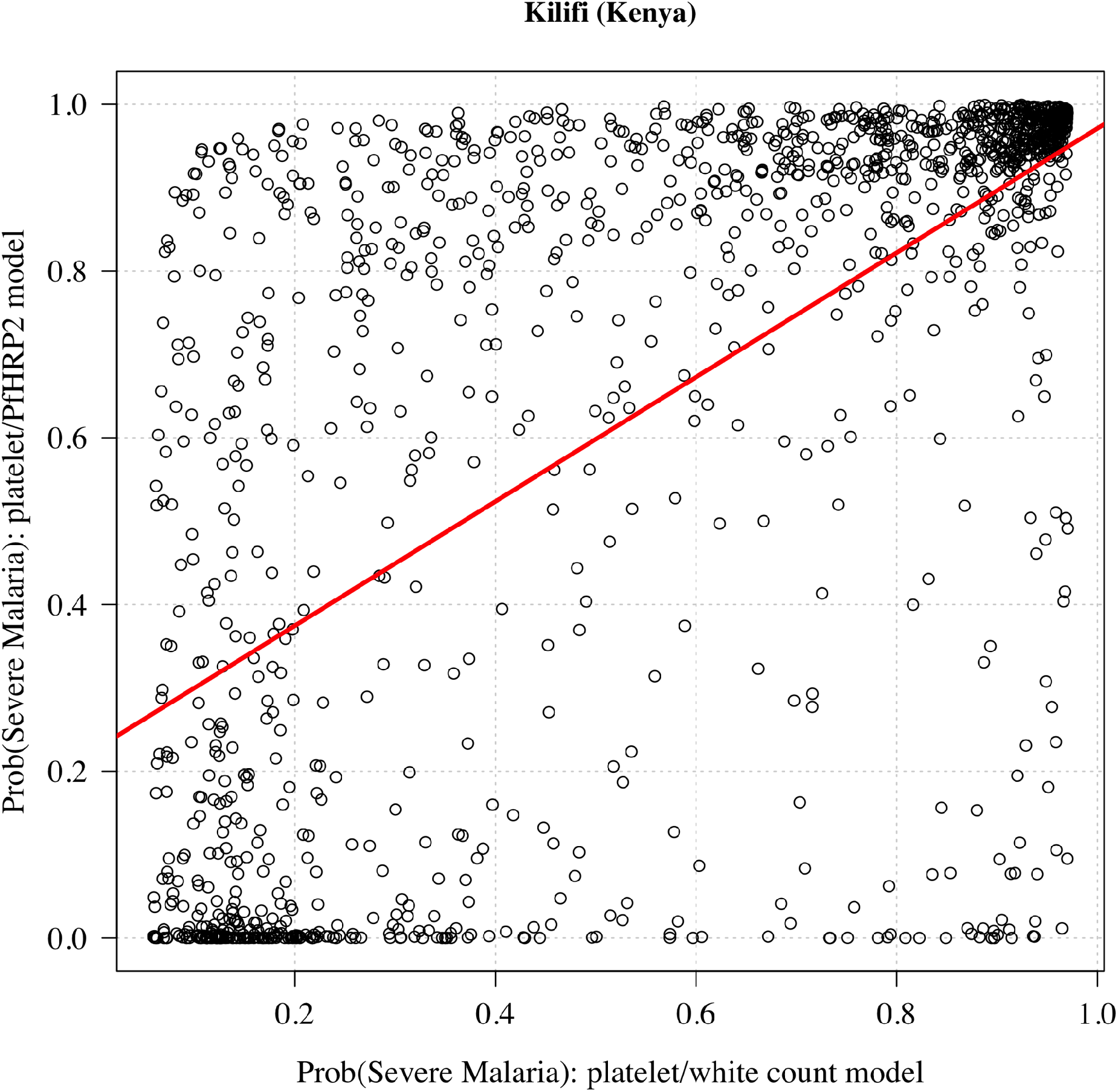
Estimated probabilties of severe malaria under the previously published platelet/white count model [4] and those under the platelet/HRP2 model for 1,400 Kenyan children with HRP2 measurements and platelet counts.

**Figure S3:**
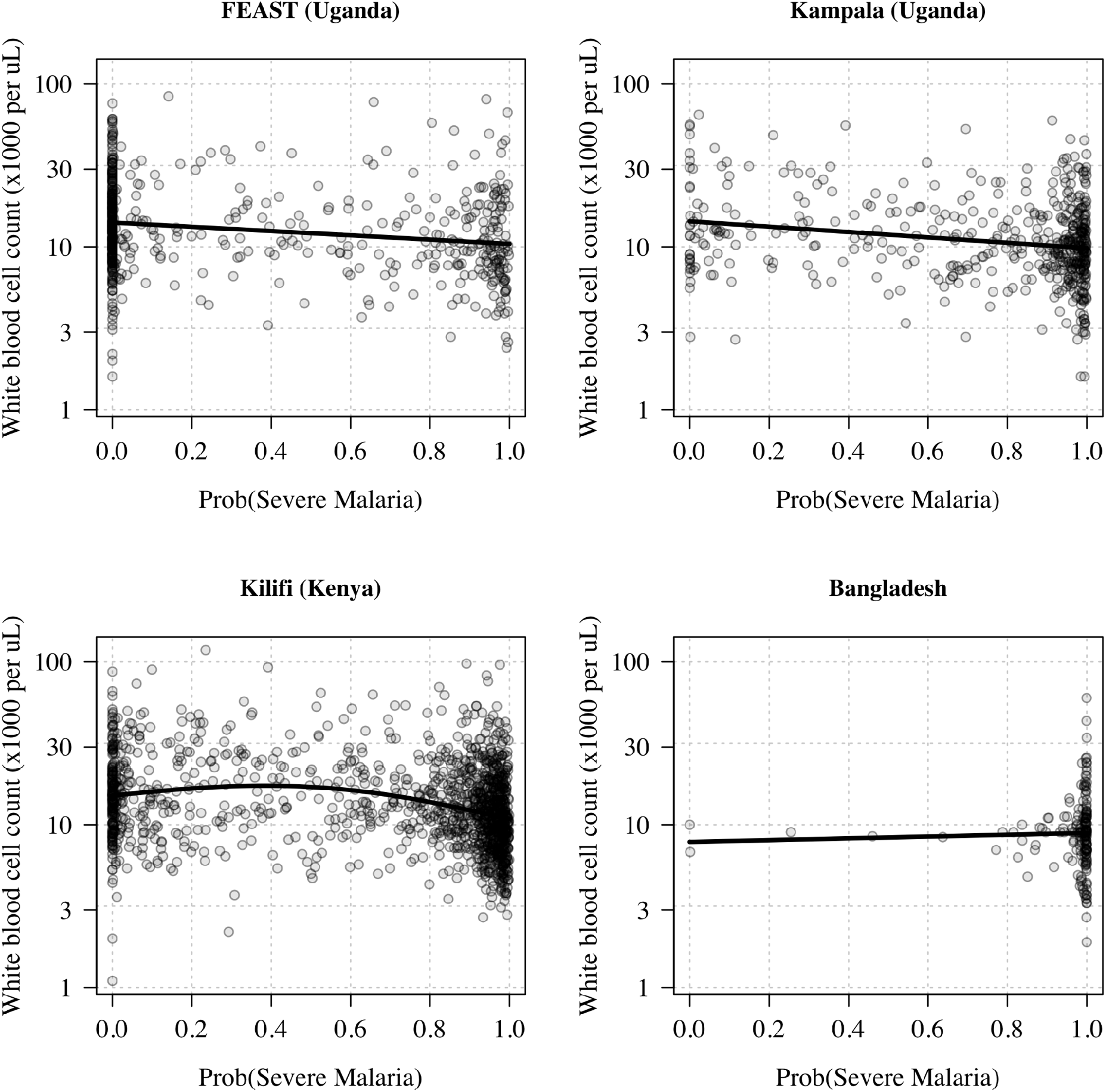
Total white blood cell counts (logarithmic scale) as a function of the model estimated probability of severe malaria (based on platelet counts and *Pf* HRP2 concentrations). The thick black lines show the overall trends.

**Figure S4:**
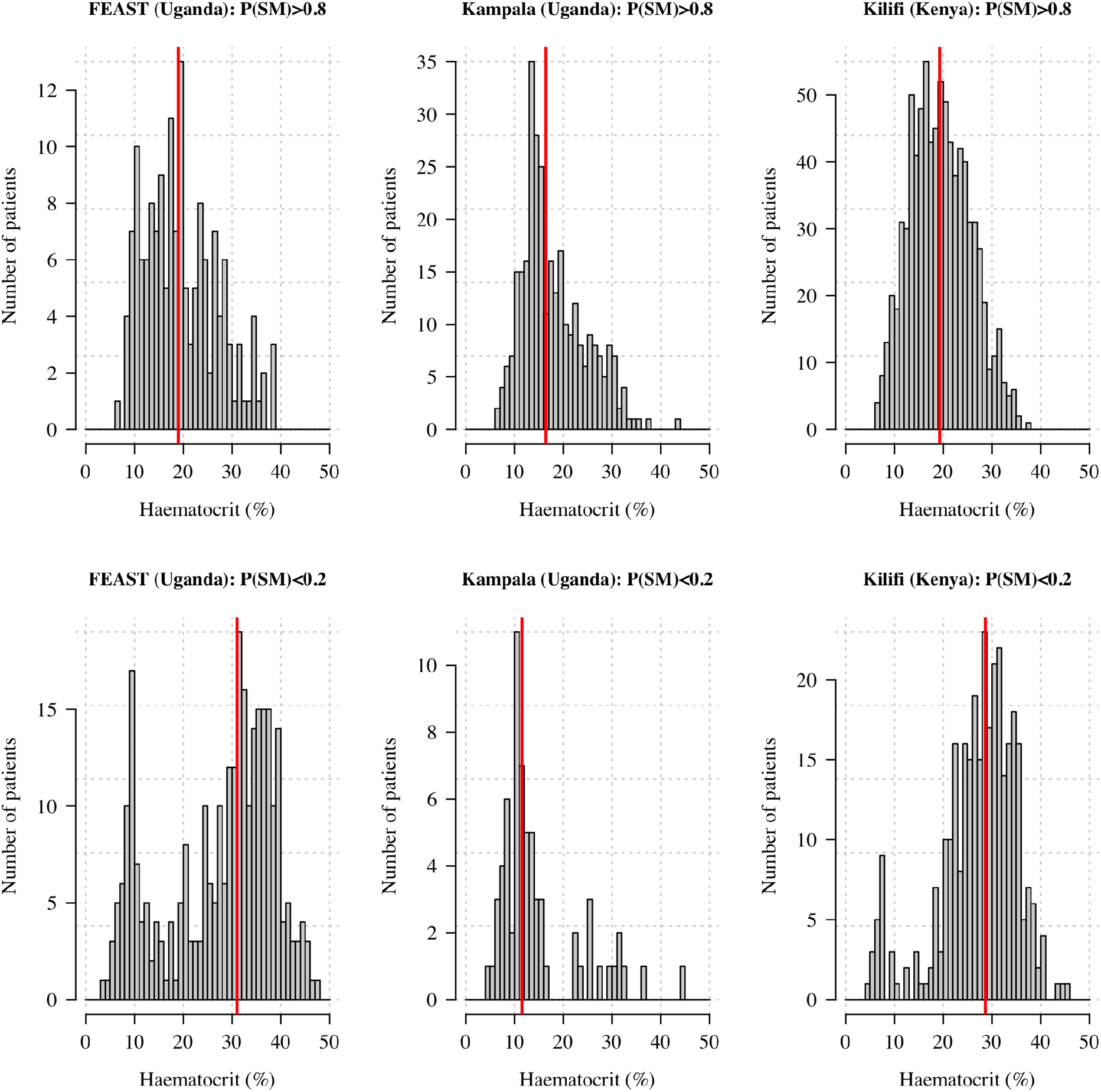
Distribution of haematocrits in African children with a high (> 0.8) and low (< 0.2) probability of having severe malaria. The vertical red lines show the median values.

